# Biomimetic Virus-like Particles as SARS-CoV-2 Positive Controls for RT-PCR Diagnostics

**DOI:** 10.1101/2020.10.16.20213991

**Authors:** Soo Khim Chan, Pinyi Du, Karole Ignacio, Sanjay Metha, Isabel G. Newton, Nicole F. Steinmetz

## Abstract

Coronavirus disease 2019 (COVID-19) is a highly transmissible disease that has affected more than 90% of the countries worldwide. At least 17 million individuals have been infected, and some countries are still battling first or second waves of the pandemic. Nucleic acid tests, especially reverse-transcription polymerase chain reaction (RT-PCR), have become the workhorse for early detection of COVID-19 infection. Positive controls for the molecular assays have been developed to validate each test and to provide high accuracy. However, most available positive controls require cold-chain distribution and cannot serve as full-process control. To overcome these shortcomings, we report the production of biomimetic virus-like particles (VLPs) as SARS-CoV-2 positive controls. A SARS-CoV-2 detection module for RT-PCR was encapsidated into VLPs from a bacteriophage and a plant virus. The chimeric VLPs were obtained either by *in vivo* reconstitution and co-expression of the target detection module and coat proteins or by *in vitro* assembly of purified detection module RNA sequences and coat proteins. These VLP-based positive controls mimic SARS-CoV-2 packaged RNA while being non-infectious. Most importantly, we demonstrated that the positive controls are scalable, stable, and can serve broadly as controls, from RNA extraction to PCR in clinical settings.

**Table of contents graphic:** 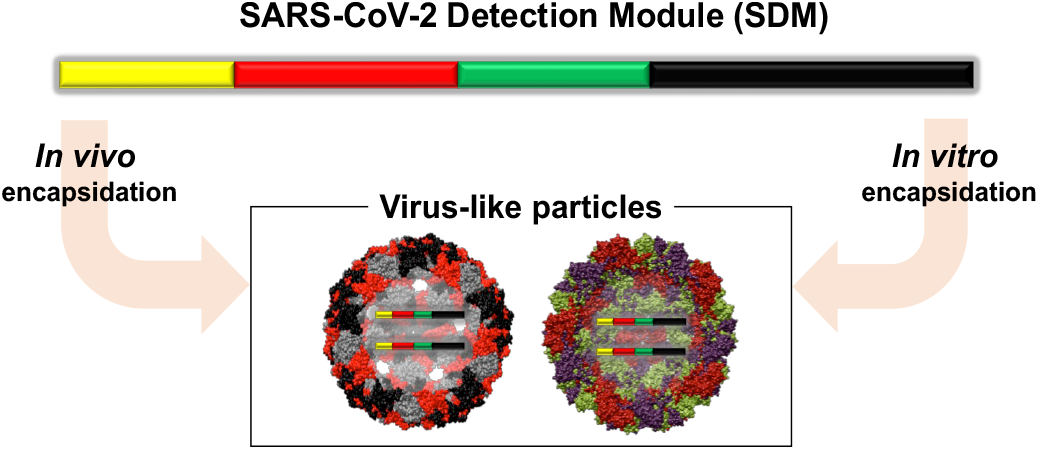

---

The ongoing coronavirus disease 2019 (COVID-19) pandemic, caused by severe acute respiratory syndrome coronavirus 2 (SARS-CoV-2), was first detected in Wuhan city, China in December 2019. As of August 8, 2020, more than 19 million confirmed COVID-19 cases have been reported globally; the United States accounted for more than a quarter of the total confirmed cases and over 161,000 deaths (https://coronavirus.jhu.edu/map.html). SARS-CoV-2 demonstrates high transmissibility during the early phase of infection due to active shedding in the upper respiratory tract and droplet transmission. Accurate diagnosis of COVID-19 cannot be made clinically alone, as it exhibits clinical manifestations common to other respiratory illnesses. ^1,2^ Moreover, asymptomatic transmission has been linked to 40-45% of COVID-19 cases. ^3^ Finally, COVID-19 disproportionately affects disadvantaged populations including minorities and people from lower socioeconomic classes where resources for testing are limited.^4^ Hence, the deployment of large-scale, rapid diagnostic testing is critical for widespread surveillance as well as early detection of infected individuals to mitigate the disease.

Quantitative reverse transcription polymerase chain reaction (RT-qPCR) is currently the gold standard for detection of SARS-CoV-2 nucleic acid owing to its high specificity, high sensitivity, and rapid turnover.^5^ Multiple sample types are being collected from patients for testing, including nasopharyngeal swabs and saliva.^6^ In order to meet the need for high capacity testing, molecular diagnostic tests such as CRISPR-based detection (Cepheid Sherlock Biosciences) and isothermal amplification technologies (ID NOW COVID-19, iAMP^®^) have recently been granted Emergency Use Authorization (EUA) by the Food and Drug Administration (FDA).^7^ Testing is the key to confine cases and end the spread of SARS-CoV-2. The World Health Organization (WHO) advised that the positivity rate (*i*.*e*. the percent of all tests that are positive) should remain at 5% or lower for at least 2 weeks before reopening.^8^ Under economic and political pressures, most localities have proceeded with erratic, phased reopenings even though the US positivity rate is about 2-fold higher as of 8^th^ August 2020.^9^ Experts agree that durable containment of SARS-CoV-2 depends on more widespread testing.^10^

Several RT-qPCR primer/probe sets that target distinct regions of the viral genome have been developed worldwide for SARS-CoV-2 detection. Testing for the nucleocapsid phosphoprotein (N) yields the most consistent test results, as it is highly conserved.^11^ The comparative sensitivity and efficiency of the different primer/probe sets were recently reviewed.^12^ The primer/probe sets developed by the Centers for Disease Control and Prevention (CDC) target two regions in the N gene: N1 at gene location 28287-28358 nucleotides (nts) and N2 at gene location 29164-29230 nts. The Human RNase P gene at gene location 28-92 nts serves as an endogenous control. Overall, these primer/probe sets are validated and yield consistent results with clinical samples.^12^ However, it remains challenging to reliably identify false negative results.

Validation of diagnostic tests is crucial to ensure test accuracy and precision. Positive controls should be run in parallel with patient samples as a benchmark to monitor and validate test accuracy. The lack of reliable positive controls is a key problem for RT-PCR-based viral testing.^13^ While some labs use infectious viral RNA extracted from highly positive patient samples, the most commonly used controls are commercially available synthetic RNA transcripts or plasmids.^14,15^ However, these samples have shortcomings. Because RNA is an inherently unstable molecule, both infectious viral RNA and synthetic RNA transcripts are labile and susceptible to degradation. Also, the multiple freeze-thaw cycles to which these samples are subjected can further degrade RNA quality.^16^ Whereas RNA stabilizers such as Paxgene^®^, RNA*later*,^17^ and DNA/RNA Shield™^18^ or production of RNA in dry form^19^ have improved the stability of RNA, the requirement for cold-chain distribution remains expensive and challenging, especially in underserved areas or regions with tropical climates. While plasmid DNA is more stable than RNA, neither positive controls enables concurrent processing with patient samples, or *full-process control*. Because RNA/DNA samples are only added downstream in the testing process, they cannot control for whether the viral RNA was degraded during the initial extraction steps.

To overcome these shortcomings, we propose a biomimetic nanotechnology solution, i.e. to package RNA transcripts containing the nucleic acid regions for binding of the SARS-CoV-2 primers and probes into a nanoparticle carrier. More specifically, we generated a biomimetic positive control by developing a virus-like particle (VLP) technology that takes advantage of the unique features of SARS-CoV-2 but is non-infectious and safe to use in diagnostic assays. While many nanotechnology platform technologies are available, including polymer and lipid nanoparticles, that could carry nucleic acids, the capsids from viruses naturally evolved to package nucleic acid and thus make a suitable choice. Specifically, biomimicry was achieved by packaging non-infectious, replication-deficient synthetic SARS-CoV-2 RNA target sequences into non-infectious VLPs from the bacteriophage Qbeta (Qβ) and the plant virus cowpea chlorotic mottle virus (CCMV). Both VLPs measure ∼30 nm and have been extensively studied in nanotechnology.^20^ Encapsidation into a viral capsid more closely mimics the conditions encountered by the RNA template of SARS-CoV-2 within clinical or environmental samples. This approach has been reported previously: for example, cowpea mosaic virus (CPMV) capsids were used to encapsidate target RNA for detection of foot-and-mouth disease virus (FMDV),^21^ tobacco mosaic virus (TMV) was developed as a positive control for Ebola diagnostic assays,^22^ and bacteriophage Qβ was explored as a control for foodborne virus detection.^23^ In response to COVID-19, Asuragen and SeraCare have announced developments of SARS-CoV-2 positive controls in which RNA targets is encapsidated into bacteriophage VLPs^24^ or a replication-deficient mammalian virus.^25^ Of note is also the development of VLPs for vaccine development;^26-27^ research from academia and industry has led to development of more than 200 vaccine candidates in record time, many design rely of nanotechnology concepts and various VLP subunit vaccines are being developed. ^28,29^

We developed a synthetic SARS-CoV-2 Detection Module (SDM). The positive controls contain synthetic SDM that contains all nucleic acid regions for binding of the CDC-designated RT-qPCR primers and probes. The SDM module was then was encapsidated into VLPs by *in vitro* or *in vivo* reconstitution of chimeric VLPs, yielding three SARS-CoV-2 positive controls termed Qβ 1P-C19, Qβ 2P-C19 (synthesized from a one or two plasmid system, respectively), and CCMV-C19. While *in vitro* reconstitution of viral capsids around a synthetic RNA template is well established (and was the method used for CCMV), the *in vivo* reconstitution is less frequently reported. However, the latter requires fewer processing steps and thus may be higher yielding and more economic to mass produce. To achieve efficient *in vivo* reconstitution, careful design considerations need to be taken in account, such as length of the target RNA and its molecular features, i.e. deletion of ribosome binding sites (for enhanced safety) and appendage of Qβ hairpin (for encapsidation).^30^ Here we developed such methods for the Qβ VLP system. The structural integrity and stability of the resulting nanoparticle positive controls were assessed over time using a combination of transmission electron microscopy (TEM), size exclusion chromatography (SEC), dynamic light scattering (DLS) and electrophoretic mobility assays. A set of RT-PCR assays was performed to validate the biomimetic SARS-CoV-2 positive controls. Lastly, the positive controls were subjected to screening and used as external controls for patient sample testing.

## RESULTS AND DISCUSSION

### Rationale for the Selection of Qβ CCMV VLPs for the Construction of COVID-19 Diagnostic Positive Controls

We chose to develop VLPs from bacteriophage Qβ and plant virus CCMV as biomimetic nanotechnology for use as positive control probes for COVID-19 diagnostic assays. Qβ and CCMV offer several advantages over current approaches that use either MS2 VLPs (Asuragen)^24^ and replication-defective mammalian virus (SeraCare).^31^ Firstly, the Qβ capsid is 5Å larger and contains approximately 20% more genomic RNA compared to MS2.^32^ Therefore, Qβ can accommodate a higher payload; and indeed Qβ can package 100 x copies of target RNA compared to MS2.^23^ Furthermore, despite extensive sequence identity between Qβ and MS2, the coat protein subunits of Qβ offer higher thermal stability compared to MS2; the Qβ capsid gains thermal stability based on inter-subunit disulfide bonds.^33^ Previous studies comparing Qβ and MS2 with comparable RNA cargo have shown that Qβ is more stable over a range of temperatures (−20°C, 4°C, and 45°C).^23^ Taking together, data suggest that Qβ would be a better candidate for long term storage or ambient shipping.

To the best of our knowledge, CCMV has never been reported for development and application of positive control in molecular diagnostic assays. A particular advantage of the CCMV system is the straightforward *in vitro* reconstitution – hence offering a high degree of modularity. Purified coat proteins could be stored and reconstituted around a target RNA cassette as needed; for example, if mutants or new strains or emerge or to adapt the probe for use in other diagnostic assays. CCMV coat proteins could be obtained through heterologous expression ^34^ or through molecular farming in plants. The latter offers a high degree of scalability and speed, and can be implemented with relatively non-sophisticated infrastructure while keeping manufacturing costs low.^35^

Furthermore, the bacteriophage and plant VLPs (Qβ and CCMV) offer advantages compared to a replication-deficient mammalian virus, which always carries a risk of residual activity or reversion, which may pose safety risks.^36-37^ Qβ and CCMV are non-infectious to mammals and unable to replicate in mammalian cells;^38^ therefore offering another layer of safety in particular for use in low-resource settings where sterile and biosafety facilities are not always attainable. Moreover, production of Qβ and CCMV through fermentation or molecular farming is more cost effective and higher yielding compared to manufacture of mammalian virus in mammalian cell cultures.^39^

### Design of the SARS-CoV-2 Detection Module (SDM)

The design of the SDM was based on the CDC-recommended detection regions. The 622 nt SDM consists of 4 segments with the first being a Qβ hairpin loop. This 29-nt Qβ hairpin has high affinity for the Qβ coat proteins^40^ and is appended to the targeted RNA sequences to facilitate SDM encapsidation into Qβ VLPs. In previous work, the Qβ hairpin loop has been used to direct the encapsidation of protein cargos such as enzymes for biocatalytic applications^41^ as well as target RNAs.^30,42^ The Qβ hairpin loop is followed by the 3 target regions: two SARS-CoV-2 N regions (Accession NC_045512.2 N1: gene location: 28271-28443; N2: gene location: 29091-29230) and the human RNase P region (or RP; Accession NM_006413: gene location: 1-280). The module is flanked by a T7 promoter and T7 terminator at 5’ and 3’ end, respectively, for RNA transcription to enable both *in vitro* and *in vivo* reconstitution of chimeric VLPs (**Figure 1** and **S1**). SDM flanked with T7 promoter and T7 terminator (SDM + T7_P/T_) was cloned into pCDFDuet™-Qβ and pET-28a (+) to generate plasmid Qβ 1P-C19 and Qβ 2P-C19, respectively (**Figure S2**). Plasmid Qβ 1P-C19 allows co-expression of the Qβ coat protein (CP) gene and SDM RNA from the same vector whereas plasmid Qβ 2P-C19 expresses only SDM RNA. pCDFDuet™-Qβ, which expresses the Qβ CP gene, was co-transformed into the same bacterial cell with plasmid Qβ 2P-C19 for *in vivo* encapsidation of SDM RNAs. The two-plasmid system has been used successfully in previous work for *in vivo* reconstitution of Qβ around target RNAs.^30,42^

**Figure 1.**
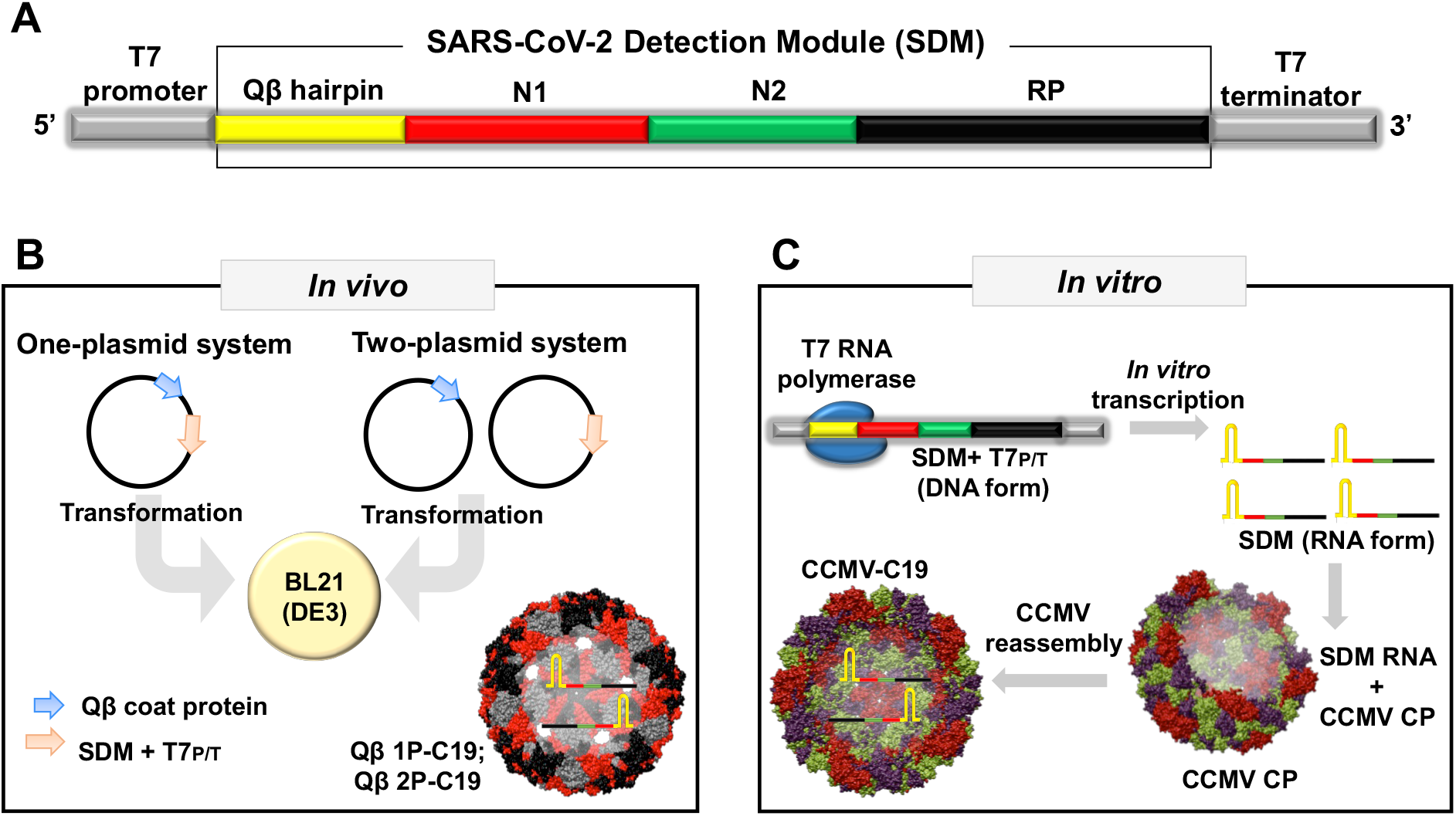
Overall schematic and workflow of the VLP-based biomimetic SARS-CoV-2 positive controls. (A) Design of SDM from 5’ to 3’: T7 promoter (grey), Qβ hairpin (yellow), N1 (red), N2 (green), and RP (black), T7 terminator (grey). (B) Production of Qβ 1P-C19 and Qβ 2P-C19 VLPs via *in vivo* assembly. Qβ 1P-C19 VLPs were produced from a one-plasmid system, where the gene of the Qβ CPs and SDM RNAs were cloned in one vector (pCDFDuet™-1). Qβ 2P-C19 VLPs were produced from a two-plasmid system, where Qβ CP was derived from pCDFDuet™-Qβ and SDM RNA was produced from pET-28a (+). (C) CCMV-C19 VLPs were produced by *in vitro* reconstitution of the transcribed SDM RNAs with purified CCMV CPs.

Several safety measures were built into the synthesis of these positive controls. SDM + T7_P/T_ was cloned out-of-frame with the open reading frame to avoid protein translation of truncated SARS-CoV-2 N1 and N2 gene segments. The ribosome binding site (RBS) upstream of SDM was removed for the same reasons. Furthermore, the presence of RBS upstream of target RNA has been shown to reduce packaging efficiency of RNAs into VLPs due to competitive binding of ribosomes versus Qβ CPs to the target RNA.^30^ Therefore, for the one- and two-plasmid systems, the RBS was removed upstream of the SDM, but RBS was retained upstream of the Qβ CPs to enable protein translation. *In vitro* transcription of SDM RNAs yielded approximately 150 µg of SDM RNAs per reaction; denaturing urea polyacrylamide gel electrophoresis of the transcribed SDM RNA confirmed the ∼622 nt RNA product (**Figure S3A**). Lastly, we used RT-qPCR assays to confirm that the SDM provided a template for the CDC primer/probe sets (**Figure S3C**).

### Production of VLP-based SARS-CoV-2 positive controls

*In vivo* encapsidation of SDM RNAs in Qβ VLPs was achieved by co-expression of Qβ CPs and SDM RNAs in *E. coli* using the aforementioned one-plasmid system and two-plasmid system to produce Qβ 1P-C19 VLPs and Qβ 2P-C19 VLPs, respectively. Production of Qβ 2P-C19 VLPs using the two-plasmid system was performed by transforming plasmid pCDFDuet™-Qβ and plasmid Qβ 2P-C19 into the same bacterial cells for co-expression of Qβ CPs and SDM RNAs. We also developed the one-plasmid system Qβ 1P-C19 from a single plasmid using the pCDFDuet™-1 vector.

For CCMV-C19, native CCMV was obtained from infected black-eyed pea No. 5 plants and disassembled to obtain purified CPs. The purified CPs were then reassembled with *in vitro* transcribed SDM RNAs (**Figure 1B**). Disassembly and reassembly are achieved through careful adjustment of the buffer conditions (see methods section). The negatively charged SDM RNAs interact with the CCMV CP, specifically with the highly positively-charged, arginine-rich binding domains at the N-terminus (amino acids 9-19; sequence: TRAQRRAAARK);^43,44^ thus reconstitution of the chimeric CCMV VLP with the packaged SDM is achieved through electrostatic interactions rather than sequence-specific interactions, as was exploited for the Qβ system.

The following SARS-CoV-2 positive controls were generated: Qβ 1P-C19, Qβ 2P-C19, and CCMV-C19. Expression of the *in vivo* reconstituted Qβ samples yielded 100 mg/L per batch of culture for Qβ 1P-C19 and Qβ 2P-C19. For CCMV, 100 g of CCMV-infected leaves yielded approximately 40 mg of CCMV nanoparticles. About 20 mg of CCMV VLPs were obtained after disassembly and about 10 mg of SDM encapsidated CCMV-C19 were obtained.

Characterization of the VLP-based SARS-CoV-2 RT-PCR positive controls revealed production of intact, pure, and monodisperse chimeric VLPs (**Figure 2**). Separation of intact VLPs on native agarose gels indicated successful RNA encapsidation, as the RNA and protein co-migrate, yielding overlapping and discrete bands when stained with GelRed™ (RNA stain) and Coomassie blue (protein stain). The VLPs have an overall negative charge and therefore migrate toward the anode.^45,46^ The size of the VLPs was determined by dynamic light scattering (DLS) and transmission electron microscopy (TEM). DLS of Qβ 1P-C19 and Qβ 2P-C19 revealed the presence of monodisperse nanoparticles with an average diameter of ∼32 nm (PD ∼0.12-0.13). For CCMV-C19, DLS also revealed monodisperse nanoparticles with a hydrodynamic radius of ∼33 nm (PDI 0.128). In all cases, broken particles, free CPs, or aggregates were not detected. These findings are in agreement with TEM imaging showing intact and uniform VLPs with Qβ measuring ∼ 26-27 nm and CCMV measuring ∼29 nm (**Figure 2**); the differences between DLS and TEM – specifically, the increased size as measured by DLS is explained by the hydration status of the sample; while ‘dry’ samples are imaged in TEM, DLS reports the hydrodynamic particle size of VLPs in solution.^47^ Size-exclusion chromatography (SEC) for all positive controls showed typical elution profiles for VLPs, where nucleic acid (260 nm) and protein (280 nm) were eluted concurrently at around 12 mL from a Superose 6 column.^46,48^ Also, the SEC results confirmed the structural integrity of particles, showing no free RNAs or CPs in the VLP preparations. Lastly, the particle properties of the chimeric VLPs packaging the SDM matched data of Qβ VLPs devoid of the SDM (**Figure S4**) as well as native CCMV particles (**Figure S5**).

**Figure 2.**
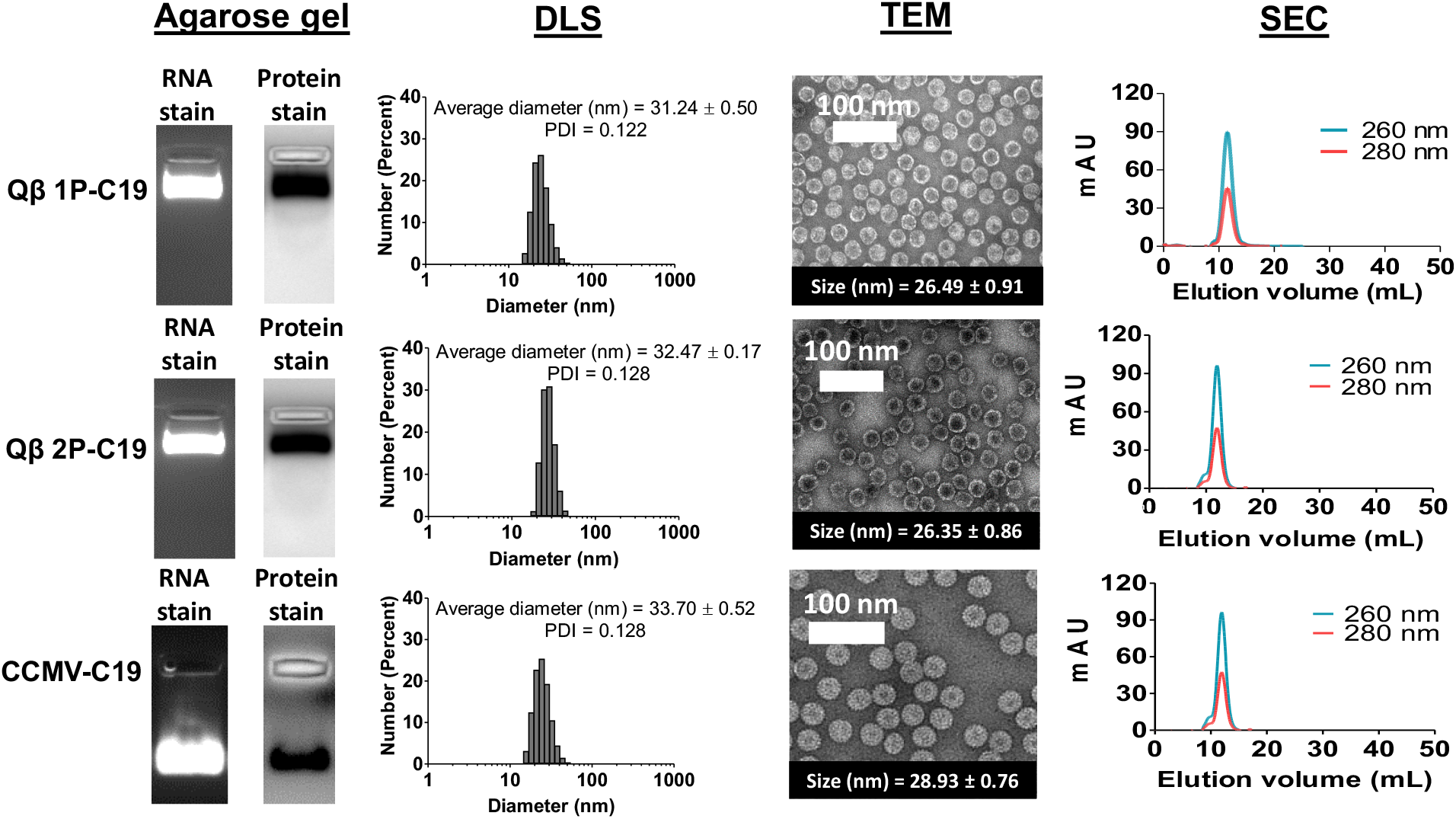
Characterization of VLP-based SARS-CoV-2 positive controls. Agarose gel: Native agarose gel electrophoresis of VLPs packaging the SDM with gels were stained with GelRed™ Nucleic Acid Stain (RNA stain), and Coomassie blue (protein stain) to show the presence of RNAs and VLPs. DLS: Dynamic light scattering (DLS) of VLPs packaging the SDM; triplicate samples were analyzed, and representative data sets are shown. TEM: Imaging of negatively stained VLPs packaging the SDM using transmission electron microscope (TEM). Average size of 20 particles tabulated by ImageJ software was stated in inset box. SEC: Analysis of positive controls by size exclusion chromatography (SEC) using a Superose 6 column and GE Healthcare Äkta Purifier chromatography system; protein was detected at 280 nm and RNA was detected at 260 nm.

### Validation of the VLP-based SARS-CoV-2 positive controls

As a benchmark, the encapsidated RNAs within the positive controls were extracted using QIAGEN QIAamp Viral RNA mini kit; the procedure followed CDC recommendations. One difference to note between SARS-CoV-2 and our control probes is that SARS-CoV-2 is an enveloped virus,^49^ but our control probes based of Qβ and CCMV are non-enveloped viruses. Nevertheless, the extraction protocol is highly suitable for RNA extraction from viruses and the lysis buffer (Buffer AVL) used in this study has been optimized by the manufacturer to isolate RNA from a wide variety of viruses, enveloped and non-enveloped. Moreover, QIAGEN QIAamp® Viral RNA Mini Kit (RNA isolation kit used in this study) is recommended by CDC and has previously been reported to isolate RNA from both enveloped and non-enveloped viruses.^50-51^ For these reasons, the structural difference between SARS-CoV-2 and the VLP control probes should not affect the results of the diagnostic assay. RNAs were extracted and then eluted in RNase-free water instead of the CDC-recommended AVE buffer for quantification and purity check. Sodium azide in the AVE buffer interferes with absorbance readings between 220 and 280 nm and was thus avoided for the quality control studies. Total nucleic acids extracted include carrier RNAs (poly A, as per manufacturer’s instructions), the target SDM RNAs, and, in the case of Qβ, also random host RNAs that are packed *in vivo*. Here, we assume that the amount of carrier RNAs is constant for each sample. When Qβ 1P-C19 and Qβ 2P-C19 were compared to Qβ VLPs, Qβ 1P-C19 and Qβ 2P-C19 packaged ∼30% more total nucleic acids compared to Qβ VLPs devoid of the SDM (**Table 1**). The higher nucleic acid content for the designer VLPs Qβ 1P-C19 and Qβ 2P-C19 may be attributed to the *in vivo* transcribed SDM RNAs with the Qβ hairpin loop. While Qβ can package RNA non-selectively based on electrostatic interactions of negatively charged cellular RNAs to the positively charged EF-loop on Qβ coat proteins,^52^ the 29-nt Qβ hairpin loop added to the SDM confers high affinity for Qβ CP^40,53^ and therefore enhances packaging and encapsidation efficiency of the SDM vs. random host RNA. While there was no significant difference in the mean of total nucleic acids extracted from Qβ 1P-C19 and Qβ 2P-C19, Qβ 2P-C19 showed higher batch-to-batch variability with the amount of total nucleic acid extracted from Qβ 2P-C19 ranging from 230 ng/µL to 360 ng/µL. This indicates that the two-plasmid system may be less reproducible and that the one-plasmid system would be the preferred system based on the higher quality control and assurance provided. Extracted RNA was analyzed by denaturing urea polyacrylamide gel electrophoresis and staining of the nucleic acids and imaging under UV light revealed presence of the SDM (622 nt). In addition, background RNAs were observed, which can be attributed to the packaging of host RNA as described above (**Figure S6A**, Lane 1-3). CCMV-C19 showed the lowest amount of total nucleic acids encapsidated, or about 50% of the total RNA extracted from Qβ positive controls. However, in stark contrast to Qβ, where the SDM only makes a fraction of the total RNA packaged (**Table 1**), for CCMV-C19, 100% of the RNA encapsidated into CCMV-C19 is the target SDM RNA (**Figure S6A**) – this is an advantage of the *in vitro* assembly system. Based on BCA assay and RiboGreen nucleic acid assay (**Table S3**), about 3 SDM RNA molecules were packaged per CCMV-C19; and this is consistent with previous reports, e.g. reporting that 4 copies of 500-nt long RNAs could be encapsidated per CCMV.^54^ In contrast, one SDM RNA molecule was encapsidated in every 17^th^ Qβ 1P-C19 particle and 30^th^ Qβ 2P-C19 particle; the inefficiency in packaging can be explained by competition of the SDM target cargo with host *E. coli* RNAs.

**Table 1.**
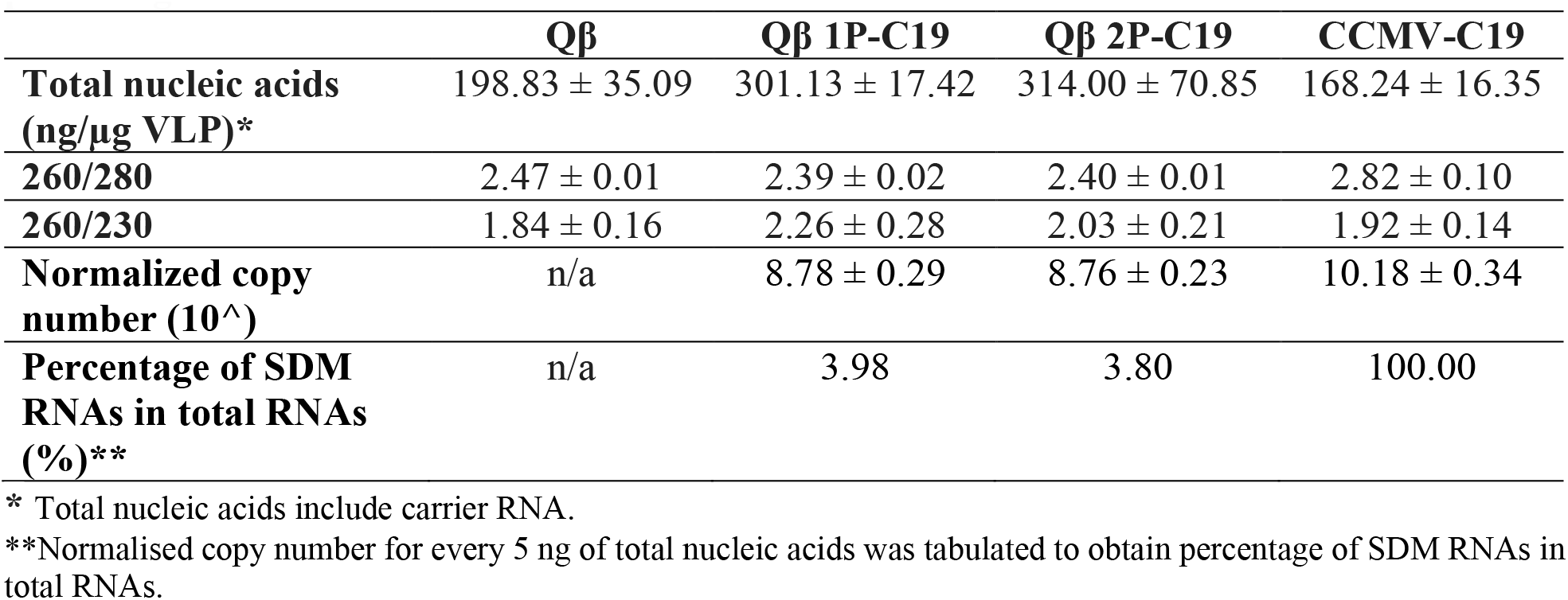
Total nucleic acids extracted from VLP-based SARS-CoV-2 positive controls and percentage of SDM.

The encapsulation of SDM into VLPs was further validated by investigating the release kinetics of SDM from the Qβ and CCMV-based probes. VLPs (Qβ 1P-C19, Qβ 2P-C19, CCMV-C19) were heated at 75°C and sampled; first samples were analyzed by agarose gel electrophoresis; RNA bands were then extracted and subjected to RT-qPCR analysis; a time course study was performed and released RNA was samples over a 5-min time course. Stable encapsulation is confirmed at t=0 mins with the protein and RNA stain co-localizing after electrophoretic separation (**Figure 3**). Upon heating of the samples, released RNA was also apparent and the intensity increased with increasing incubation time. The heating also reduced the mobility of the coat proteins, which further suggest disassembly and RNA release.^55,56^

**Figure 3.**
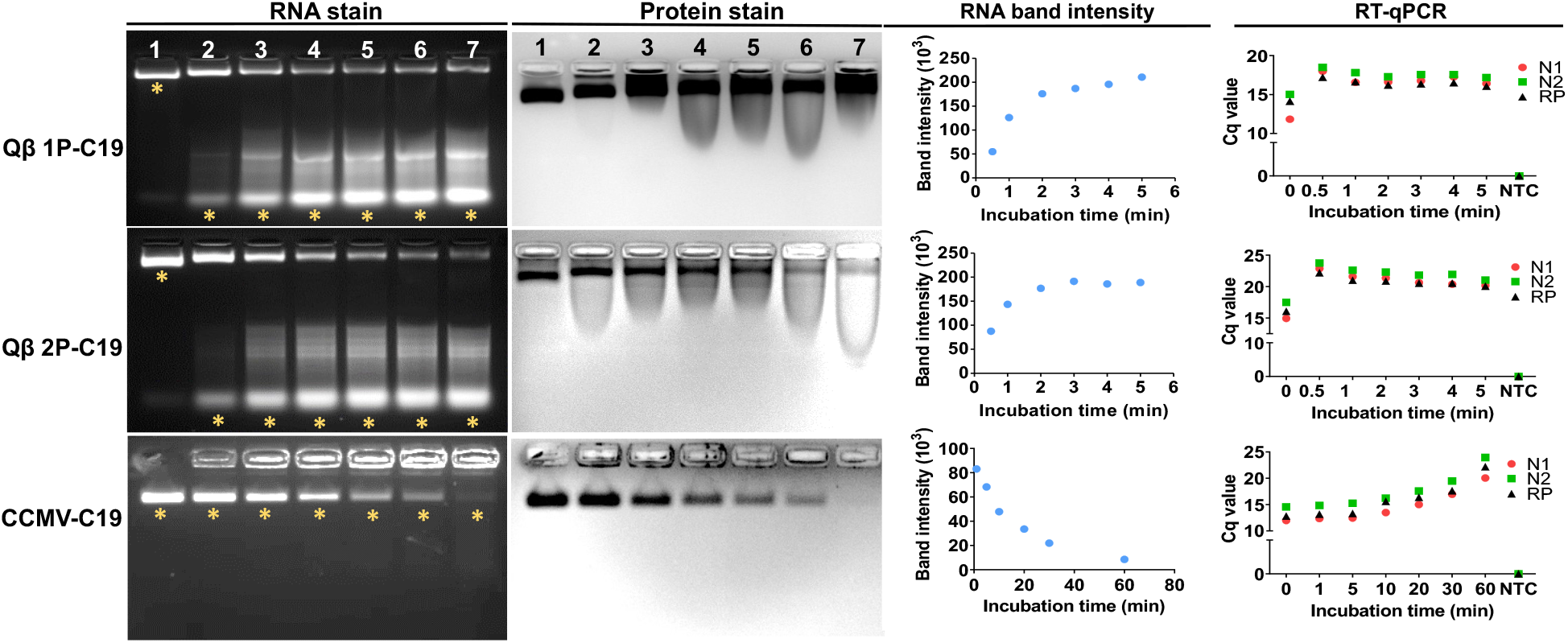
Release kinetics of encapsidated SDM RNAs from VLPs at 75°C at various time points as determined by agarose gel electrophoresis and RT-qPCR. VLPs were analyzed by native agarose gel electrophoresis after heating at 75°C for 0 min (Lane 1), 0.5 min (Lane 2), 1 min (Lane 3), 2 min (Lane 4), 3 min (Lane 5), 4 min (Lane 6), and 5 min (Lane 7) for Qβ 1P-C19 and Qβ 2P-C19. CCMV-C19 was heated at 75°C for 0 min (Lane 1), 1 min (Lane 2), 5 min (Lane 3), 10 min (Lane 4), 20 min (Lane 5), 30 min (Lane 6), and 60 min (Lane 7). Same gels were stained with GelRed™ Nucleic Acid Stain (RNA stain), and Coomassie blue (protein stain). RNA bands excised from gels for RT-qPCR were labelled by yellow asterisks. RNA band intensities from 30 s were evaluated by ImageJ software. RT-qPCR was performed using all three sets of primer/probe sets (N1, N2, RP). NTC represents no-template control in RT-qPCR.

To confirm that SDM were indeed encapsidated into the VLPs, RNAs from time point 0 (intact particles) was extracted from gels and validated by RT-qPCR. Presence of N1, N2, and RP fragments was confirmed (**Figure 3**; RT-qPCR panel). We also carried out RT-qPCR analysis on RNA released from the Qβ 1P-C19 and Qβ 2P-C19 VLPs upon heating at 75°C and presence of SDM RNAs and its N1, N2, and RP was confirmed. Similar observations were made for the CCMV-C19 construct; however, to achieve effective release longer incubation was needed to release the RNA. Also, for the CCMV-C19 probe, RT-qPCR confirmed presence of N1, N2, and RP, the components of the SDM RNA. The amount of SDM RNAs quantified by RT-qPCR matched the RNA band intensity measured. It was interesting to note that RNA release kinetics were slower for faster for Qβ vs. CCMV, because Qβ possess higher thermal stability.^57^ The porous structure of Qβ likely is the underlying reason for the faster RNA release kinetics observed.

Amplification efficiency is one of the most important factors in qPCR. An ideal amplification efficiency of 100% corresponds to exponential doubling of the PCR product during every cycle to give an amplification factor of 2.^58^ The slope of standard curve is be used to tabulate the amplification efficiency, with the equation: E = (10^−1/slope^ – 1) x 100%.^59^ Amplification efficiency of the CDC primer/probe set has been validated with synthetic SARS-CoV-2 RNA transcripts as well as clinical samples.^12,60^ N1 and N2 primer sets have reported amplification efficiencies of more than 90%, which validates that the primers are optimized for SARS-CoV-2 RNA binding. Here, we confirmed the amplification efficiency of the CDC primers on our SDM RNA. All three regions (N1, N2, RP) showed amplification efficiency above 90%, with correlation coefficient (R^2^) >0.99 (**Figure 4**). N1 and RP primer/probe sets demonstrated higher sensitivity as compared to N2 primer/probe set and enabled detection of our SARS-CoV-2 positive controls at 10^1^ copies/µL for Cq <40. For the N2 primer/probe set, the detection limit was 10^2^ copies/µL. This result aligned with a previous report when using real patient samples,^12^ indicating that our SARS-CoV-2 positive controls are able to function as a SARS-CoV-2 mimic for RT-qPCR detection.

**Figure 4.**
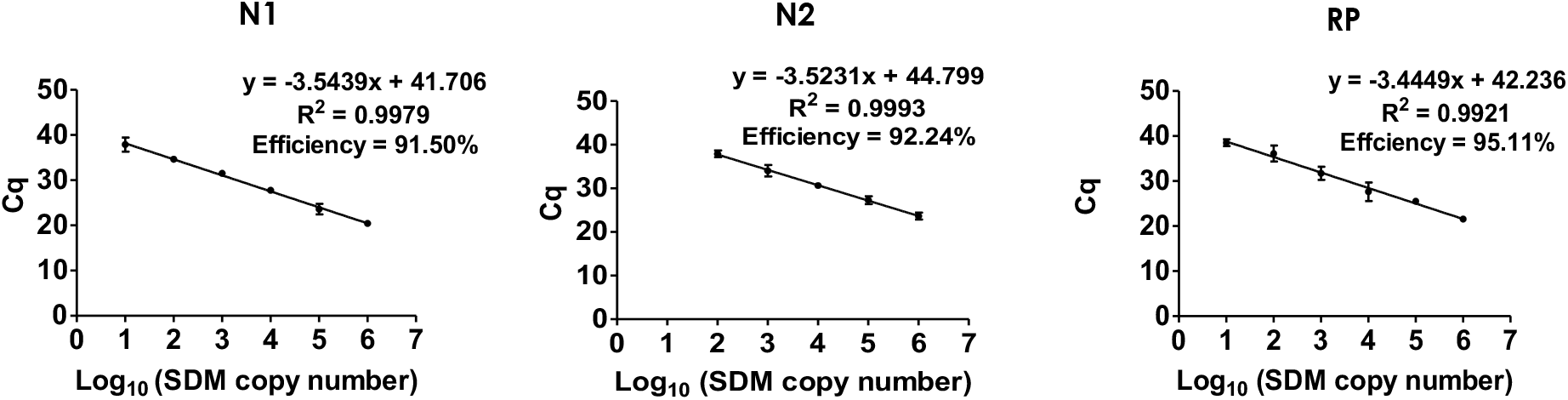
Validation of CDC primer/probe sets on *in vitro* transcribed SDM RNAs. RT-qPCR was performed on *in vitro* transcribed SDM RNAs (10^6^-10^0^ copies) to construct N1, N2, and RP standard curves for tabulating PCR amplification efficiency. Error bars show the standard deviation.

Using the RT-qPCR method, we determined the SDM RNA copy number per nanogram of total nucleic acids extracted (after normalization of the Cq value, see **Figure S6B**). The SDM RNA copy number was tabulated from mean copy number of N1, N2, and RP based on the standard curves shown in **Figure 4**. Proper baseline settings are critical for accuracy.^61^ Therefore, to standardize the baseline setting in RT-qPCR, we first normalized the quantification cycle (Cq) value of CCMV-C19 to *in vitro* transcribed SDM RNA by assuming that total RNAs extracted from CCMV-C19 contain only SDM RNA. The normalized ratio is then applied to Cq values of Qβ 1P-C19 and Qβ 2P-C19, as mentioned in **Figure S6B**, to obtain the percentage of SDM RNAs in total RNA extracted. CCMV-C19 has the highest SDM RNA copy number, with >10^9^ copies per nanogram of total nucleic acids (**Table 1**). This is as expected, because CCMV-C19 was assembled *in vitro* using only SDM target RNAs. Qβ 1P-C19 and Qβ 2P-C19 have similar copy number of SDM RNA, or around 10^8^ for every nanogram of total nucleic acids (**Table 1**). The lower copy number is owing to encapsidation of cellular *E. coli* RNAs, which has been shown in **Figure S6A**. All CDC primer/probe sets demonstrated no binding to random cellular *E. coli* RNAs that encapsidated in Qβ.

There was no significant difference in SDM RNA copy number between Qβ 1P-C19 and Qβ 2P-C19 (unpaired t-test, p > 0.05), suggesting that the one-plasmid system with indirect upstream RBS has similar packaging efficiency as the two-plasmid system (though the two-plasmid system resulted in greater batch-to-batch variability, as discussed above). The SDM RNAs made up about 4% of the total RNAs encapsidated in Qβ. Despite the target RNAs only being a fraction of the total RNAs extracted, the copy number of encapsidated SDM RNAs is still sufficiently high to function as a positive control. Early infection yields about 6.76 × 10^5^ copies per whole swab.^1^ At least 10^8^ copies of SDM RNAs were obtained from every microgram of VLP (similarly to every 5 ng of total nucleic acids extracted). Compared to other VLPs proposed as SARS-CoV-2 positive controls, the Qβ VLPs offer higher production efficiency, yielding VLPs at packing with about 100-fold mores copies of RNA compared to, for example, MS2 bacteriophage.^23^

### Validation of the VLP-based SARS-CoV-2 positive controls in the clinical setting

In-house VLP-based SARS-CoV-2 positive controls were validated in clinical settings by comparing performance alongside with clinical samples using a droplet digital PCR system (ddPCR). Clinical testing revealed that the VLP-based SARS-CoV-2 positive controls yielded amplitude signals between 2000 to 10,000 for all the three regions (N1, N2, RP) (**Figure 5A**). The 1-D amplitude plot confirmed our in-house VLP-based SARS-CoV-2 positive controls presents a functional SARS-CoV-2 RNA mimic and enables detection using the CDC primer/probe set (**Figure 5A**). All three detection regions recommended by CDC (N1, N2, RP) in all three SARS-CoV-2 positive controls, Qβ 1P-C19, Qβ 2P-C19, and CCMV-C19, were successfully detected. Importantly, it was possible to process the positive controls alongside the clinical samples throughout every step, starting from RNA extraction though amplification, providing seamless full-process control.

**Figure 5.**
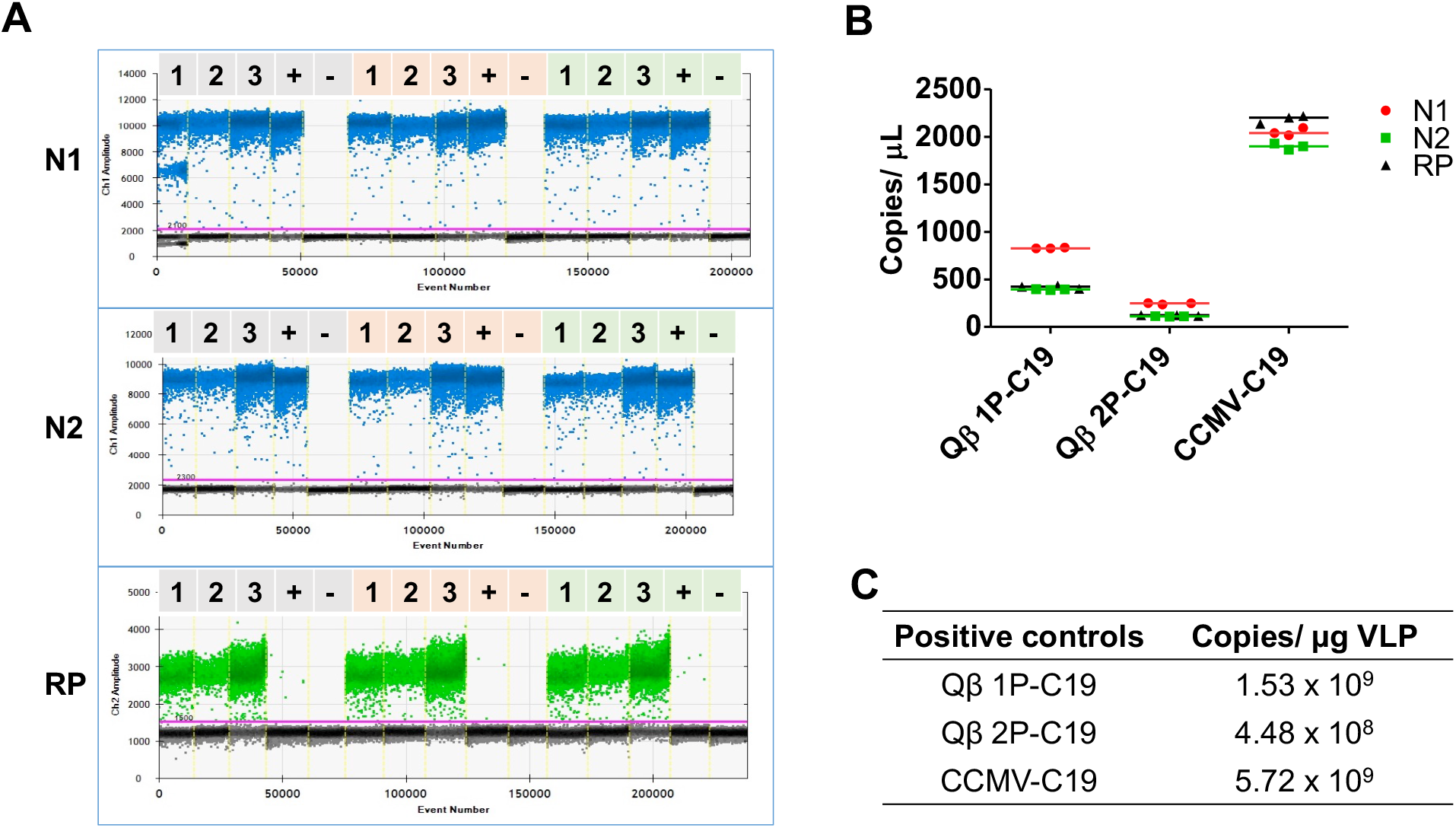
Validation of VLP-based SARS-CoV-2 positive controls in the clinical setting using ddPCR detection of SARS-CoV-2. (A) ddPCR 1-D amplitude plots of SARS-CoV-2 positive controls according to N1, N2, and RP regions. Lane 1: Qβ 1P-C19. Lane 2: Qβ 2P-C19. Lane 3: CCMV-C19. Lane 4: Clinical sample from COVID-19 confirmed patient. Lane 5: Clinical sample from healthy patient for N1 and N2 (negative control); no template control for RP. Amplifications were performed in triplicate. (B) Scatter plot comparing copy numbers of SARS-CoV-2 detection regions (N1, N2, RP) for all positive controls. (C) Tabulated SDM RNA copy number for each SARS-CoV-2 positive controls.

Consistent with the above-described findings, CCMV-C19 probes were the most sensitive, as reflected by the high copy numbers detected (> 2,000 copies/µL, **Figure 5B**); this is consistent with CCMV packaging the highest copy number of SDM. Fewer copies of SDM were packaged in Qβ 1P-C19 and Qβ 2P-C19 (due to encapsidation of random host RNAs) and this resulted in lower SDM copies detected (<1,000 copies/µL and <500 copies/µL, respectively, **Figure 5B**). Overall, the data were aligned with the results obtained from RT-qPCR (**Table 2**).

**Table 2.**
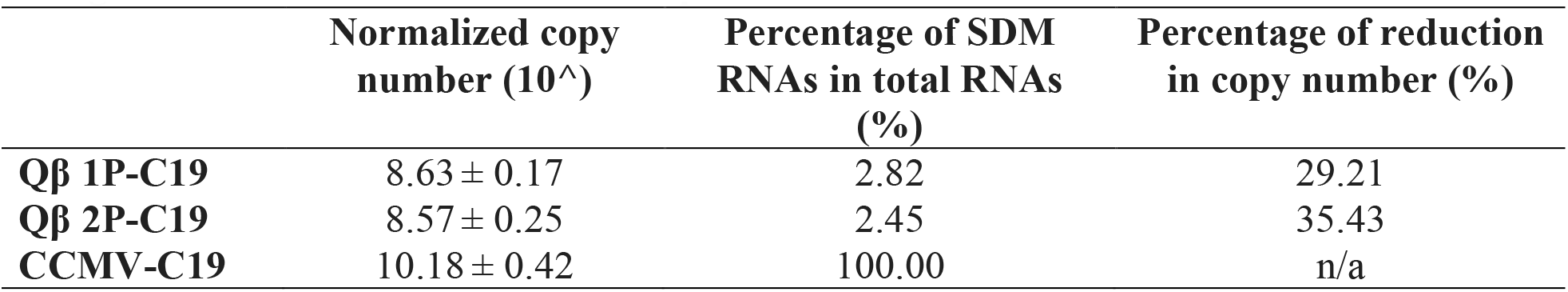
Normalized copy number of SDM per 5 ng of total nucleic acids after 1-month storage of the VLP-based SARS-CoV-2 positive controls.

When comparing the performance of Qβ 1P-C19 vs. Qβ 2P-C19, it was interesting to note the differences comparing ddPCR vs. RT-qPCR: while RT-qPCR showed no apparent differences in copy number of SDM (**Table 1**), ddPCR indicated higher SDM copy for Qβ 1P-C19 *vs*. Qβ 2P-C19, with 1.53 ⨯10^9^ SDM/µg VLP *vs*. 4.48 ⨯10^8^ SDM/µg VLP (**Figure 5C**). The introduction of PCR inhibitors at the RNA extraction stage may account for the reduced sensitivity of the RT-qPCR method.^62^ Previous studies have shown that ddPCR method exhibits higher tolerance to PCR inhibitors compared to conventional RT-qPCR and this is explained by reaction partitioning.^63,64^ Also, droplet-based reaction partitioning in ddPCR will lead to increased variance in SDM copy number when comparing the positive controls. Regardless of the differences in performance observed, each of the proposed SARS-CoV-2 positive controls - Qβ 1P-C19, Qβ 2P-C19, and CCMV-C19 - provided robust positive controls with encapsidate at least 10^8^ SDM copies from every microgram of VLP (**Figure 5C**).

### Stability of the VLP-based SARS-CoV-2 positive controls

Most of the commercially available positive controls require cold-chain distribution in order to maintain product quality. However, the pipeline of cold-chain distribution starting from packaging to logistics increases the cost of the product and limits their widespread distribution. Cold-chain infrastructure is not available in low-resource settings and is especially problematic in countries with tropical climates. Bacteriophages and plant viruses evolved as highly stable capsids that retain their structural properties under various environmental conditions, making them suitable candidates for development of positive controls that do not require a cold-chain infrastructure. To investigate the stability of our in-house SARS-CoV-2 positive controls under conditions that simulate shipping conditions, samples were left at room temperature (approximately 20-25°C) for a month. Structural integrity and SDM functionality were then assessed using agarose gel electrophoresis, DLS, TEM, and SEC, as well as RT-qPCR. Qβ 1P-C19 and Qβ 2P-C19 exposed to these conditions remained intact. There was no aggregation and there were no free CPs to indicate disassembly of the Qβ positive controls, and the particle characteristics matched those obtained of freshly prepared particles (**Figure 6** vs. **Figure 2**). The excellent stability of Qβ can be attributed to the two cysteine residues on the Qβ coat protein forming inter-subunit disulfide bonds.^53^

**Figure 6.**
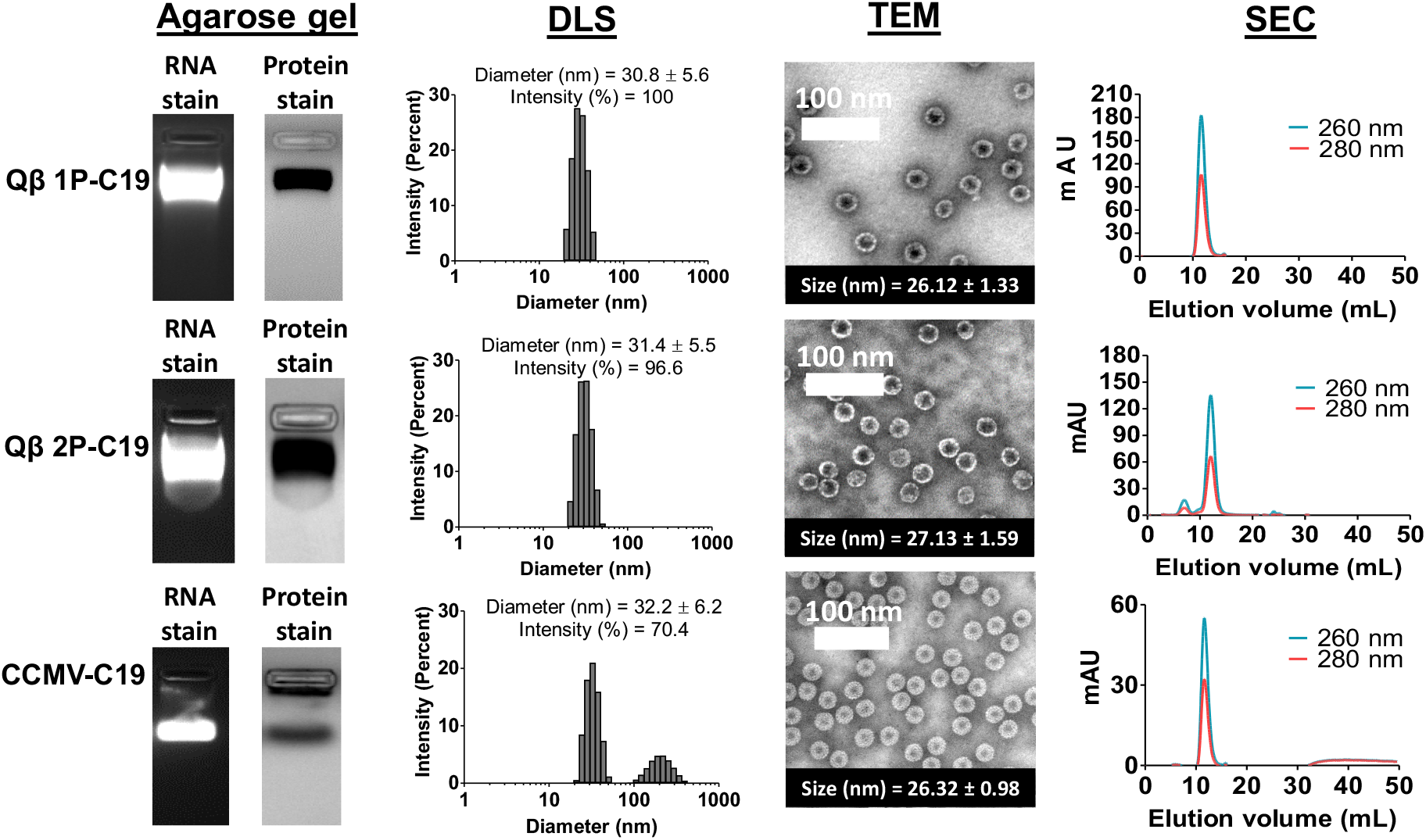
Characterization of VLP-based SARS-CoV-2 positive controls after 1-month storage under ambient conditions. Agarose gel: Native agarose gel electrophoresis of VLP-based SARS-CoV-2 positive controls reveals intact VLPs; gels were stained with GelRed™ Nucleic Acid Stain (RNA stain), and Coomassie blue (protein stain) to show the presence of RNAs and VLPs. DLS: Dynamic light scattering (DLS) of VLPs packaging the SDM; triplicate samples were analyzed, and representative data sets are shown. TEM: Imaging of negatively stained VLPs packaging the SDM using transmission electron microscope (TEM). Average size of 20 particles tabulated by ImageJ software was stated in inset box. SEC: Analysis of positive controls by size exclusion chromatography (SEC) using a Superose 6 column and GE Healthcare Äkta Purifier chromatography system; protein was detected at 280 nm and RNA was detected at 260 nm. See also **Figure 2**; showing the characterization of freshly-prepared samples.

**Figure 7.**
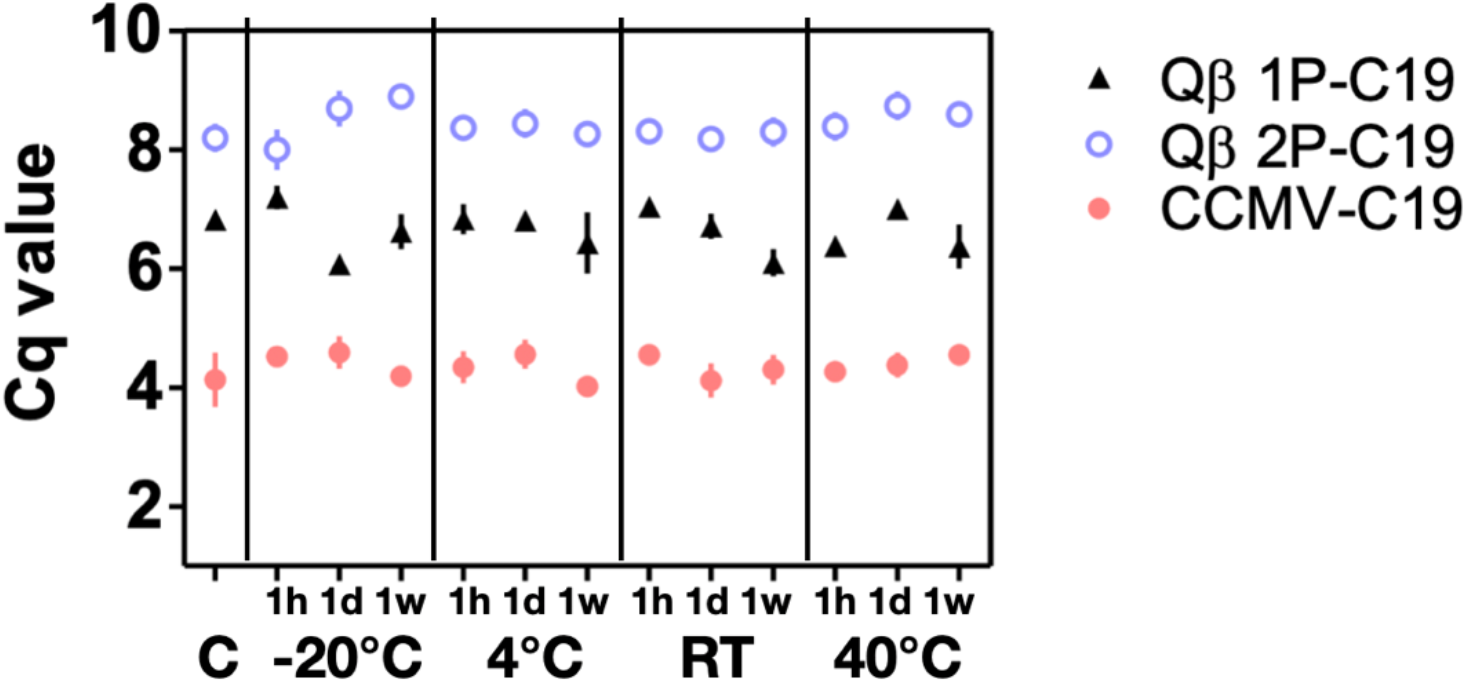
Stability of VLP-based SARS-CoV-2 positive controls in respect to time and temperature. RT-qPCR was performed to obtain the Cq values. Triplicates were performed on each sample with error bar shows the standard deviation.

The CCMV-C19 positive control was found to be less stable, however still yielding a recovery of ∼70% intact CCMV-C19 positive control after 1-month storage at ambient conditions (**Figure 6**, DLS panel). The CCMV assembly is stabilized by electrostatic and hydrophobic interactions.^65,66^ However, the lack of disulfide bonds in CCMV reduces longitudinal particle stability.^67^ Data are consistent and indicate partial disassembly leading to some degree of aggregation: native gel electrophoresis revealed intact particles with RNA and CPs having matched electrophoretic mobility; however a second protein band appeared near the well, indicating aggregation of CPs and RNA (**Figure 6**, Lane 2). While the RNA stain was less obvious, this pattern may indicate a mixture of disassembled CPs associated and/or aggregated with RNA via their positively-charged N-terminal tails.^68^ VLP aggregation was not apparent from TEM imaging, therefore we attribute the agarose gel electrophoretic pattern as well as the observed aggregates in the DLS measurement to partial disassembly of CPs and their aggregation as CP-RNA complexes. Indeed, SEC of CCMV-C19 indicates a second broadened elution peak at 35-50 mLs. For application in the clinic, it may be desired to stabilize the CCMV capsid through introduction of covalent bonds between CPs; this concept was recently demonstrated using a homobifunctional cross-linker 3, 3’-dithiobis (sulfosuccinimidylpropionate) (DTSSP) to form intra-particle cross-links between lysine side chains of adjacent CPs.^43^

Next, and most importantly, we assayed for SDM stability. While no structural changes where observed for the Qβ 1P-C19 and Qβ 2P-C19 positive controls, RT-qPCR data indicate that the SDM copy number was reduced by 30-35% (**Table 2**). This observation is aligned with the reduction of total nucleic acid extracted from one-month-old positive controls (**Table S1**). We deduced the reduction of RNAs is due to hydrolysis contributed by water molecules in buffered solution providing hydroxyl or hydronium ions for proton transfer.^69^ The Qβ capsid is a porous structure and has pores measuring 1.3-1.4 nm at its 3-fold axis as well as 0.7 nm-sized pores at its 5-fold axis. These pores facilitate diffusion of water (causing RNA hydrolysis), ions, as well as degraded RNAs (leading to loss of RNA content).^32,55^ A possible strategy to stabilize the Qβ positive control for future applications may be to plug the pores through appropriate chemistry or through freeze-drying in appropriate excipients.^70^

Lastly, an expected reduction in SDM copy number was also apparent for CCMV-C19. A 30% reduction of SDM was estimated from the dissociation observed (**Figure 6**), and this is consistent with reduction of total nucleic acids (**Table S1**) after the 1-month storage period.

We further challenged the VLP probe stability with respect to time and temperature by incubating the three VLPs (Qβ 1P-C19, Qβ 2P-C19, and CCMV-C19) at four different temperatures (−20°C, 4°C, room temperature: 20°C-25°C, 40°C) for 1 h, 1 day, and 1 week, respectively. These conditions mimic most of the shipping and storage conditions. Data indicate no apparent reduction in Cq value which correlates to SDM RNA copy number under any of the environmental conditions tested. CCMV-C19 encapsidated the highest amount of SDM RNAs by giving the lowest overall Cq values compared to its Qβ companions. Qβ 2P-C19 has the highest overall Cq values due to encapsidation of lowest amount of SDM RNAs as mentioned previously. Cq values fluctuated within ± 0.55, ± 0.45, and ± 0.29 cycles for Qβ 1P-C19, Qβ 2P-C19, and CCMV-C19, respectively. It should be noted that fixed volume instead of fixed amount of RNAs was applied in the RT-qPCR assay, therefore variation in Cq values correlates to the amount of target SDM. CCMV-C19 showed the most consistent Cq values with ∼16 % variation in SDM RNA copy number over the various environmental conditions and time course. This may be attributed to the fact that CCMV encapsidates only the synthetic SDM RNAs; in contrast Qβ-based probes also harbor a significant amount of ‘junk’ RNAs that may impact the quality of the RT-qPCR. Moreover, the increased variation in Cq values for the Qβ 1P-C19, and Qβ 2P-C19, with ∼25 % fluctuation, may also be explained by the porous nature of the capsid which may be more prone to RNA hydrolysis.^69^ Nevertheless, overall stability of the VLP constructs is maintained and demonstrated over the 1-week time course at various temperatures; even incubation at 40°C for one week did not significantly impact the SDM packaged in the VLP probes; therefore our proposed biomimetic control probes may be suitable for use also in low-resource settings.

## CONCLUSIONS

In conclusion, both plant virus-derived CCMV and bacteriophage-derived Qβ VLPs offer promising platforms for the encapsidation of RNA modules and application as stable, widely applicable positive controls for RT-qPCR or ddPCR detection of infectious agents, such as SARS-CoV-2. We developed *in vitro* reconstitution protocols and *in vivo* expression systems, yielding CCMV- and Qβ-based nanoparticles encapsidating SARS-CoV-2 detection modules that are compatible with the CDC primer/probe sets. Because the RNA is stabilized inside the VLP particle, the positive control mimics the conditions encountered by the RNA template of SARS-CoV-2 within clinical or environmental samples. The increased stability also enables for these positive controls to be applied as full-process controls, as demonstrated in the clinical assays performed. Both, the CCMV and Qβ platforms are massively scalable through manufacture via plant molecular farming and bacterial fermentation. The *in vivo* expression of Qβ VLPs offers the advantage of fewer processing steps compared to *in vitro* dis-assembly and reassembly used to obtain the chimeric CCMV VLPs. The latter method does offer control of target molecule encapsidation: while the payload of CCMV is solely SDM (∼ 10^9^ SDM copies/µg VLP), only a fraction of the Qβ payload is SDM (∼ 10^8^ SDM copies/µg VLP), with a large portion of the cargo being non-target host RNAs. Nevertheless, both VLP systems were robust in clinical assays. Importantly, the developed positive controls are safe and avoid the risks of using RNA extracted from infected patients. The positive controls demonstrated considerable stability over 1-month at ambient conditions; further, the probes offered excellent stability at temperatures as high as 40°C over one-week (longer time periods were not tested). Together, these attributes made the handling of SARS-CoV-2 positive controls safe and accessible for clinical personnel in a wide range of settings; given the ease of manufacture and stability conferred over a range of environmental setting, the proposed designs may aid diagnostic testing not just in testing facilities, but also at the point of entry (i.e. at airports or border crossings) and in low-resource areas. The potential to make these SARS-CoV-2 positive controls widely available at ambient conditions could help to alleviate some of the disparities in testing that are contributing to the increased COVID-19-related deaths in underserved populations in the US and across the world.

## MATERIALS AND METHODS

### Construction of Qβ 1P-C19 and Qβ 2P-C19 plasmids

*Qβ 1P-C19*: SARS-CoV-2 detection module (SDM) was synthesized and cloned into pCDFDuet™-Qβ between restriction site NotI and NdeI to generate Qβ 1P-C19 plasmid (GenScript, **Figure S1**). The gene was cloned not in frame with the open reading frame (for safety reason; the placement out of open reading frame avoids translation) and placed downstream of Qβ coat protein gene.

*Qβ 2P-C19*: SARS-CoV-2 detection module (SDM) gene from plasmid Qβ 1P-C19 was subcloned into plasmid pET-28a(+) by amplifying with Qβ 2P-C19 Forward primer (5’-GAA GAT CTT AAT ACG ACT CAC TAT AGG G-3’) and Qβ 2P-C19 Reverse primer (5’-TTT TCC TTT TGC GGC CGC CAA AAA ACC CCT CAA GAC CCG TTT AGA G-3’) using NEB Q5^®^ High Fidelity 2X Master Mix. The gene was cloned between restriction site BgIII and NotI in pET-28a(+), devoid of ribosome binding site at upstream to generate plasmid Qβ 2P-C19. This plasmid was used to express SDM RNAs *in vivo*. Plasmid pCDFDuet™-Qβ was co-transformed with plasmid Qβ 2P-C19 to express Qβ coat proteins for *in vivo* reconstitution of SDM RNAs. Clone was subjected to DNA Sanger sequencing (Eurofins Genomics) to confirm the insertion.

### Production of Qβ 1P-C19 and Qβ 2P-C19 VLPs

Plasmids were transformed into BL21 (DE3) competent *E. coli* cells (New England Biolabs^®^) and plated out on antibiotic containing plate. Selection of Qβ 1P-C19 transformants was based on streptomycin resistance (100 µg/mL); while Qβ 2P-C19 transformants was based on streptomycin resistance (100 µg/mL) and kanamycin (50 µg/mL). The *E. coli* was inoculated in Luria-Bertani (LB) media supplemented with antibiotic and incubated overnight at 37°C with shaking at 250 rpm. For VLP expression, the overnight culture was diluted 1: 100 in Thermo Fisher Scientific’s MagicMedia™ *E. coli* Expression Medium and incubated overnight at 30°C with shaking at 300 rpm. The culture was centrifuged at 9800 x *g* for 10 min at 4°C. The cell pellet was resuspended with 0.1 volume of 1 X phosphate buffered saline (PBS), pH 7.4 supplemented with 0.2 mg/mL lysozyme and sonicated for 10 min with 5 sec on/off cycle. The lysate was then centrifuged at 9800 x *g* for 15 min to collect the supernatant. Crude VLPs in supernatant were pelleted down with 10% (w/v) PEG 8000 and 0.2 M NaCl followed by resuspension in 1 X PBS (pH 7.4). Crude VLPs were treated with 0.7 volumes of 1:1 chloroform: butanol mixture and the upper aqueous layer was purified on 10-40% (w/v) sucrose gradient by centrifugation at 133,000 x *g* for 3 h, 4°C. The band with VLPs was collected and centrifuged at 210,000g for 3 h, 4°C. The final pellet was resuspended in 1 X PBS (pH 7.4) and the VLPs were stored at -20°C. VLP concentration was measured with Pierce™ BCA protein assay kit.

### *In vitro* transcription of SARS-CoV-2 detection module

Plasmid Qβ 1P-C19 was digested with NotI and NdeI to obtain the linearized SDM. *In vitro* RNA transcription of SDM was performed with Thermo Fisher Scientific’s MEGAscript™ T7 Transcription kit and purified with Invitrogen™’s MEGAclear™ Transcription Clean-Up kit. Purity and concentration of transcribed RNAs was validated with Thermo Scientific™ Nanodrop 2000/2000c at ratio 260/280 and 260/230. RNA concentration is determined from A260 with reading 1.0 is equivalent to about 40 ng/µL of RNA. Pure RNA should yield around 2 or higher for both ratios. *In vitro* transcribed RNAs were also analyzed using Invitrogen™ Novex™ 6% TBE-Urea gel (cat. no: EC6865BOX).

### Production of CCMV-C19 VLPs

Primary leaves of *Vigna unguiculate*, California black-eyed peas No. 5 were mechanically infected with CCMV after growing for 12 days (these protocols are carried out under USDA-approved P526 permits). Plants were grown for another 12 days before leaves were harvested. CCMV was purified using established procedures.^72^ In brief, harvested CCMV infected leaves were homogenized with Preparation Buffer (0.2 M NaOAc, 1 mM EDTA, PH 4.8) and filtered through cheesecloth. Filtrate was later centrifuged at 15,000 x *g* for 15 min. The supernatant was precipitated by 0.02 M NaCl, and 8% (w/v) PEG 8000 and stirred overnight at 4°C. The solution was centrifuged at 15,000 x *g* for 10 min at 4°C. The pellet was resuspended in 20 mL CCMV Buffer (0.1 M NaOAc, 1 mM EDTA, PH 4.8) followed by centrifugation at 8000 x *g* for 10 min at 4°C. The supernatant was then centrifuged over a 5 mL 20% (w/v) sucrose cushion in water at 148,000 x *g* for 2 h at 4°C. The pellet was resuspended in CCMV Buffer. Purified CCMV was stored as intact virion until further use. Purified coat proteins (CPs) were obtained using established disassembly protocols.^54,73^ Disassembled CCMV CPs were reassembled with purified SDM obtained through *in vitro* transcription from plasmids; the SDM-to-CP ratio was 6:1 as previously described.^54^ The reconstituted CCMV was stored in CCMV Buffer at -80°C. VLP concentration was measured with Pierce™ BCA protein assay kit.

### Characterization of VLP-based SARS-CoV-2 Positive controls

#### Transmission electron microscopy (TEM)

Positive controls (Qβ 1P-C19, Qβ B 2P-C19, CCMV-C19) were diluted to 0.2 mg/mL in Milli-Q water and 4 µL was adsorbed to Formvar/carbon-coated 400 mesh copper grids (Electron Microscopy Science) for 2 min. The grid was washed with 4 µL of water for 2 min followed by adsorption of 4 µL of 2% (w/v) uranyl acetate (Fisher Scientific) for 2 min. Solution was removed from grid by blotting with filter paper. TEM grids were imaged with FEI Tecnai G2 Spirit transmission microscope at 80 kV. The size of particles was analyzed using ImageJ software. 20 particles were randomly selected and their diameters measured.

#### Dynamic light scattering (DLS)

Qβ 1P-C19 and Qβ 2P-C19 were diluted to 0.2 mg/mL in 1 X phosphate buffered saline (PBS, pH 7.4) Qβ 1P-C19 and CCMV-C19 samples were diluted to 0.2 mg/mL using Virus Suspension Buffer (VSB: 50 mM sodium acetate, 8 mM magnesium acetate [pH 4.5]). 60 µL samples were then analyzed with a Malvern Panalytic Zetasizer Nano ZSP.

#### Agarose gel electrophoresis

*Qβ 1P-C19 and Qβ 2P-C19*: 10 µg of Qβ 1P-C19 and Qβ 2P-C19 positive controls were loaded onto a 1.2% (w/v) TAE agarose gel. The samples were electrophoresed at 110 V for 40 min.

*CCMV-C19*: 10 µg of CCMV-C19 in a total volume of 15 µL in VSB was mixed with 3 µL of 100% glycerol before loading into 1%(w/v) agarose gel buffered with virus electrophoresis buffer (0.1 M sodium acetate, 1 mM EDTA, pH 5.5). The samples were electrophoresed at 50 V for 60 min at 4°C. Documentation of gels was performed with ProteinSimple FluorChem R.

#### Size exclusion chromatography (SEC)

200 µL of 0.5 mg/mL of VLP-based SARS-Cov-2 positive controls were analyzed by GE Healthcare Äkta Purifier chromatography system using a Superose 6 column. Samples were analyzed at a flow rate of 0.5 mL/min using 1 x PBS for Qβ 1P-C19 and Qβ 2P-C19; VSB for CCMV-C19. Detectors were set at 260 nm (RNA) and 280 nm (protein).

#### Quantification of encapsidated SDM RNA molecules in VLP-based SARS-Cov-2 positive controls

1 µL of VLP was quantified with Pierce™ BCA protein assay kit and Thermo Fisher Quant-iT™ RiboGreen™ RNA Assay Kit according to manufacturer’s protocol. Number of encapsidated SDM RNA molecule per VLP was calculated as follows:

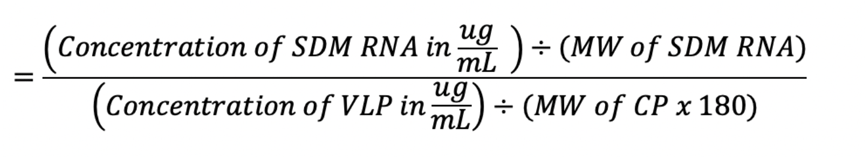

*MW SDM RNA = 199kDa; MW CCMV CP = 21kDa; MW Qβ CP = 14.3kDa

#### Extraction of total RNAs from VLP-based SARS-Cov-2 positive controls

RNA was extracted from VLP-based SARS-Cov-2 positive controls using QIAGEN’s QIAamp Viral RNA mini kit according to the manufacturer’s protocol. RNA was eluted with 50 µL of nuclease-free water. Purity and concentration of total RNAs was determined by Thermo Scientific™ Nanodrop 2000/2000c. Total RNAs were analyzed using Invitrogen’s Novex™ TBE-Urea gel, 6%. The gels were stained with 2 µL of BioGold GelRed™ Nucleic Acid Stain at 10, 000 X in 40 mL of water for 20 min and washed off with 40 mL of water for 10 min. Documentation of gels was performed with ProteinSimple FluorChem R.

### Quantitative reverse-transcription polymerase chain reaction (RT-qPCR): validation of VLP-based SARS-CoV-2 positive controls

PCR amplification efficiency was performed with a range of SDM RNA transcripts (10^6^ to 10^0^ copy) using 2019-nCoV CDC qPCR Probe Assay from Integrated DNA Technologies (IDT) (cat. no: 10006713) and Invitrogen™’s SuperScript™ III Platinum™ One-step RT-qPCR kit (cat. No: 11732020) according to the manufacturer’s protocol. Briefly, 2 µL of RNA was used in a 20 µL reaction containing final concentration of 1 x reaction mix, 0.4 µL of SuperScript™ III RT/Platinum™ Taq Mix, and 1 x primer/probes (IDT™). PCR cycling conditions were performed as followed: 50 °C for 15 min, 95 °C for 2 min, followed by 40 cycles of 95 °C for 15 seconds and 60 °C for 30 sec. The RT-qPCR reactions were performed on BioRad CFX96 Touch™ Real-Time PCR Detection System. All samples were run in triplicate. Quantification cycle (Cq) values were tabulated by CFX™ Maestro Software.

### RNA release kinetics from VLP-based SARS-CoV-2 positive controls

SARS-CoV-2 positive control (10 µg) in a total volume of 20 µL was incubated at 75°C for 0 min, 0.5 min, 1 min, 2 min, 3 min, 4 min, and 5 min. The solution was later analyzed with native agarose gel electrophoresis as mentioned previously and band intensity was analyzed by ImageJ software. The RNA bands were excised by scalpel and soaked in 500 µL of 1 x TE buffer, Molecular Biology Grade (Promega) supplemented with 40 U RNase inhibitor (Applied Biosystem™). The mixture was incubated at room temperature for 25 min with gentle shaking. The gel was later removed from TE buffer and RNA was extracted using Thermo Scientific™ GeneJET Gel Extraction Kit according to manufacturer’s protocol with slight modification. Briefly, 1 volume of Binding buffer was added to 1 volume of gel and incubated at 56°C until dissolved (6-8 min). The solution was transferred to column and spun 1 min at 14,550 x *g*. The column was washed twice with 700 µL of Wash buffer by spinning at 14,550 x *g* for 1 min. An additional spin at similar condition was performed to remove residual ethanol. Then, 30 µL of nuclease-free water was added to the center of membrane and incubated at 56°C for 5 min. Lastly the column was spun at 14,550 x *g* for 1 min to elute the RNA. 1 µL of eluted RNA was quantified with RT-qPCR using N1, N2, and RP primers/probe, respectively.

### Validation of the VLP-based SARS-CoV-2 positive controls in the clinical setting

SARS-CoV-2 positive controls (10 µg) were extracted using QIAGEN’s QIAamp Viral RNA mini kit (cat. no: 52904) according to manufacturer’s protocol and eluted in 140 µL of AVE buffer. 10 µL of eluted RNA sample was diluted to 10^−6^ with RNase-free water. Clinical samples extracted from a COVID-19 case (positive control patient) and a healthy patient (negative control). Clinical samples were diluted 100 times with RNase-free water. 10 µL of the diluted RNA sample was used to set up singleplex ddPCR in a 20 µL reaction with Bio-Rad One-step RT-ddPCR Advanced Kit (cat. no: 1864022) according to the manufacturer’s protocol. Briefly, the reaction mixture consisted of 5 µL of 4 X One-step RT-ddPCR supermix for probes, 10 µL of RNA, 2 µL of reverse transcriptase, 1 µL of 300 nM DTT, 900 of nM each forward and reverse primer and 250 of nM probe. N1, N2 and RP primer/probes were synthesized from Integrated DNA Technologies (**Table S2**). The mixture was then used for droplet generation by adding 70 μL of Bio-Rad droplet generation oil (cat. no: 1864007). Droplets were generated with Bio-Rad QX200™ Droplet Generator. The droplets were incubated at 25°C for 3 min, 45°C for 60 min, 95°C for 10 min and then cycled at 95°C for 30 sec and at 55°C (N1, N2,) or 60°C (RP) for 60 sec. Amplification was performed for 45 cycles using Applied Biosystems^®^ Veriti^®^ 96 well Thermal Cycler. The droplets from each sample were read on QX200™ droplet reader machine. The data was processed using QuantSoft™ version 1.7.4 software.

### Stability of the VLP-based SARS-CoV-2 positive controls

SARS-CoV-2 positive controls (10 µg) were aliquoted to a total volume of 20 µL and stored at four different temperature (−20°C, 4°C, room temperature: 20°C-25°C, 40°C) for 1 h, 1 day and 1 week, respectively. RNA was extracted from VLPs and 1 µL of eluted RNA was used in RT-qPCR as mentioned previously using N1 primers/probe. All samples were assayed in triplicate on BioRad CFX96 Touch™ Real-Time PCR Detection System. Quantification cycle (Cq) values were tabulated by CFX™ Maestro Software.

## Data Availability

NA

## AUTHOR INFORMATION

### Authors

**Soo Khim Chan** − *Department of NanoEngineering, University of California−San Diego, La Jolla, California 92039, United States; orcid*.*org/0000-0002-1336-2882*

**PinYi Du** – *Department of Medicine, University of California−San Diego, La Jolla, California 92039, United States*

**Karole Ignacio** – *Department of Medicine, University of California−San Diego, La Jolla, California 92039, United States*

**Sanjay Metha** – *Department of Medicine, University of California−San Diego, La Jolla, California 92039, United States*

**Isabel G. Newton** – *Department of Radiology, Division of Interventional Radiology, University of California and Veterans Administration San Diego Healthcare System −San Diego, California 92161, United States*

## Author Contributions

SKC designed the VLPs and performed the experimental work. PD and KI performed assays on patient samples. SM and IN helped in the development of the project. NSF conceived the study and oversaw the VLP design and testing. SKC and NFS wrote the manuscript; all authors read and edited the manuscript.

## Notes

The authors declare no competing financial interests.

## ACKNOWLEDGMENT

This work was funded in part by grants from the National Science Foundation: RAPID CBET-2032196 and RAPID CMMI-2027668, as well as the University of California: UCOP-R00RG2471 and a Galvanizing Engineering in Medicine (GEM) Award. Drs. Vero Beiss and Oscar Ortega-Rivera (UC San Diego) are thanked for helpful discussion regarding VLP design.

## ABBREVIATIONS

COVID-19: coronavirus disease 19;
SARS-CoV-2: severe acute respiratory syndrome coronavirus 2;
RT-qPCR: quantitative reverse-transcription polymerase chain reaction;
ddPCR: droplet digital polymerase chain reaction;
Qβ: Qbeta;
CCMV: cowpea chlorotic mottle virus;
VLP: virus-like particle;
SDM: SARS-CoV-2 Detection Module.

## Supporting Information Available

This material is available free of charge via the Internet at http://pubs.acs.org. Supporting figures (Figure S1-S6) and Supporting table (Table S1) showing nucleic acid sequences, vectors, and additional VLP characterization.

