## Supplemental Information for "Biomimetic Virus-like Particles as SARS-CoV-2 Positive Controls for RT-PCR Diagnostics"

5’- ***TAA TAC GAC TCA CTA TAG GG*** AAATGC A TGTC TAA GAC AGC AT CTTC AAAATGTCTG ATAATGGACC
CCAAAATCAG CGAAATGCAC CCCGCATTAC GTTTGGTGGA CCCTCAGATT CAACTGGCAG TAACCAGAAT
GGAGAACGCA GTGGGGCGCG ATCAAAACAA CGTCGGCCCC AAGGTTTACC CAATAATACT GCGTCTTGGT
TCACCGCTCT CAC CTTTCGGCAG ACGTGGTCCA GAACAAACCC AAGGAAATTT TGGGGACCAG GAACTAATCA
GACAAGGAAC TGATTACAAA CATTGGCCGC AAATTGCACA ATTTGCCCCC AGCGCTTCAG CGTTCTTCGG
AATGTCGCGC ATGGGACTTC AGCATGGCGG TGTTTGCAGA TTTGGACCTG CGAGCGGGTT CTGACCTGAA
GGCTCTGCGC GGACTTGTGG AGACAGCCGC TCACCTTGGC TATTCAGTTG TTGCTATCAA TCATATCGTT
GACTTTAAGG AAAAGAAACA GGAAATTGAA AAACCAGTAG CTGTTTCTGA ACTCTTCACA ACTTTGCCAA
TTGTACAGGG AAAATCAAGA CCAATTAAAA TTTTAACTAG ATTAACAATT ATTGTCTCGG ATCCATCTCA
CTGCAATGTT ***CTA GCA TAA CCC CTT GGG GCC TCT AAA CGG GTC TTG AGG GGT TTT TTG*** -3’

**Figure S1:** SARS-CoV-2 detection module (SDM) is flanked by T7 promoter and T7 terminator at 5’ and 3’, respectively, for *in vitro* transcription. Both T7 promoter and T7 terminator are italised bolded in blue. The Qβ hairpin is underlined in orange, SARS-CoV-2 N1 region is in red, SARS-CoV-2 N2 region in green and RP region in black. The SDM flanked with T7 promoter and T7 terminator (SDM + $T7_{P/T}$) was cloned not in frame with open reading frame of plasmid to avoid proper protein translation of truncated SARS-CoV-2 gene.

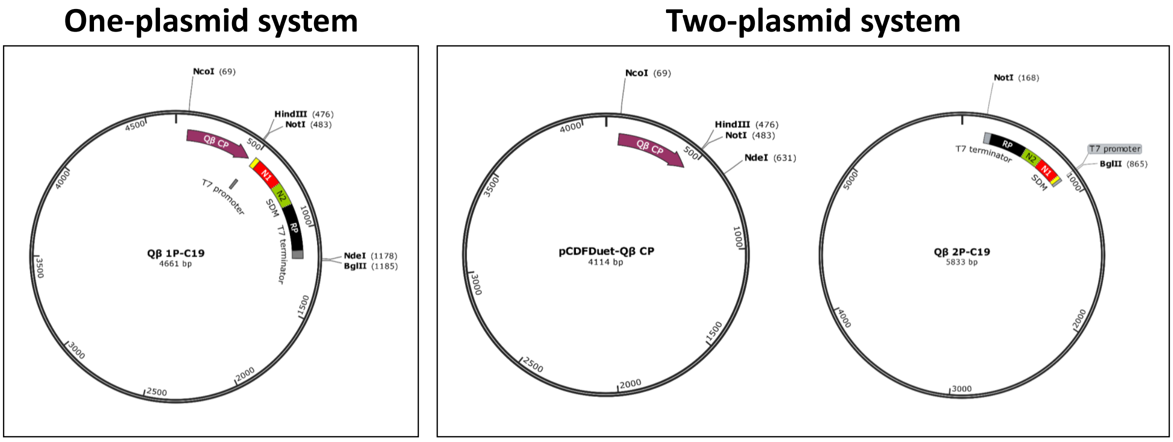


**Figure S2:** Vector maps of plasmids for production of Qβ 1P-C19 VLPs and Qβ 2P-C19 VLPs. **(Left)** One-plasmid system for production of Qβ 1P-C19 VLPs: SDM + $T7_{P/T}$ was cloned downstream of Qβ capsid protein in plasmid pCDFDuet™-Qβ to construct plasmid Qβ 1P-C19. **(Right)** Two-plasmid system for production of Qβ 2-C19 VLPs: SDM + $T7_{P/T}$ was cloned in pET-28a (+) to construct plasmid Qβ 2P-C19 and co-express Qβ capsid protein from pCDFDuet™-Qβ. N1, N2, and RP in SDM were indicated as red, green, and black, respectively.

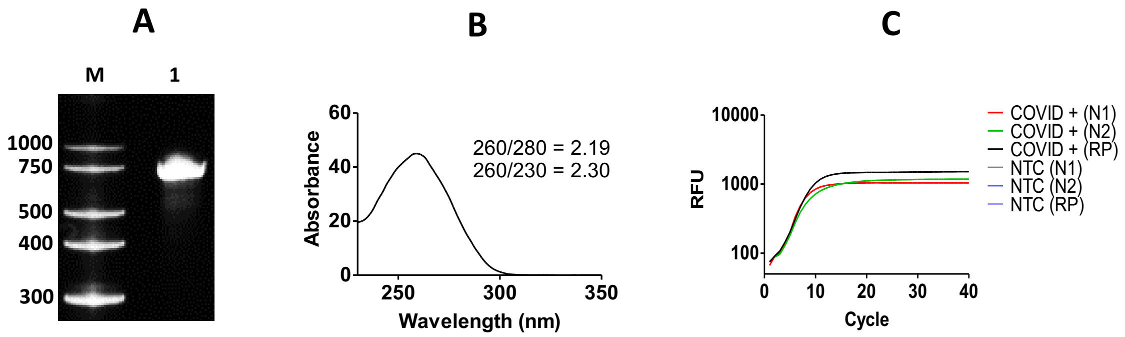


**Figure S3:** Characterization of the *in vitro* transcribed SDM RNA. **(A)** Analysis of *in vitro* transcribed SDM RNA on a denaturing 6% urea polyacrylamide gel. M: Thermo Fisher Scientific Century™-Plus RNA Markers. Lane 1: 200 ng of SDM RNA. **(B)** Determination of purity of SDM RNA by Nanodrop. Ratio 260/280 and 260/230 were determined. **(C)** RT-qPCR of *in vitro* transcribed SDM RNA (COVID +) by US CDC primers/probes sets for all the three regions (N1, N2, RP). No template control (NTC) serves as negative control.

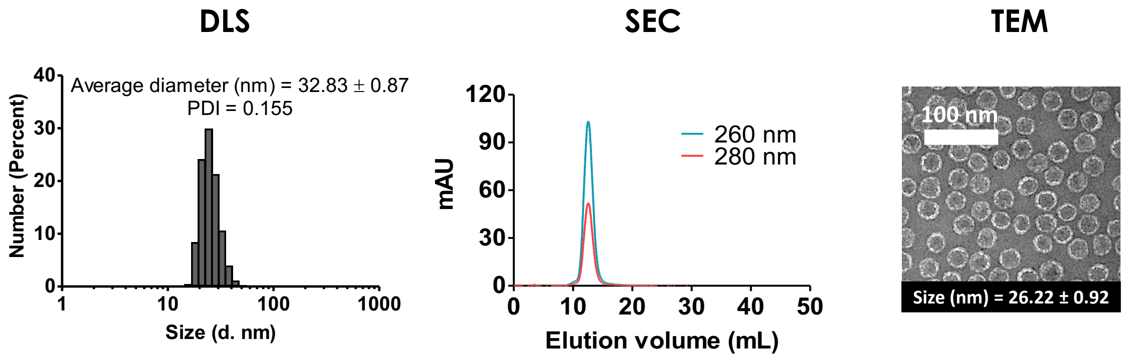


**Figure S4:** Characterization of Qβ VLP by DLS, SEC, and TEM.

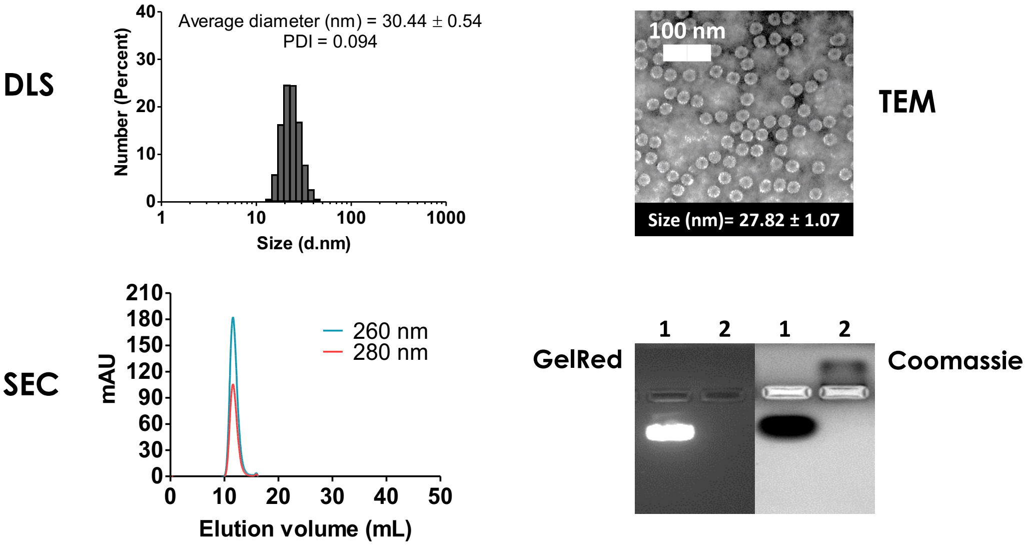


**Figure S5:** Characterization of wild type (WT) CCMV by DLS, SEC, and TEM. Disassembled CCMV was analysed by agarose gel shown at right bottom panel. Lane 1: WT CCMV. Lane 2: Disassembled CCMV. The same gel was stained by GelRed and Coomassie blue to show the presence/absence of RNA.

A

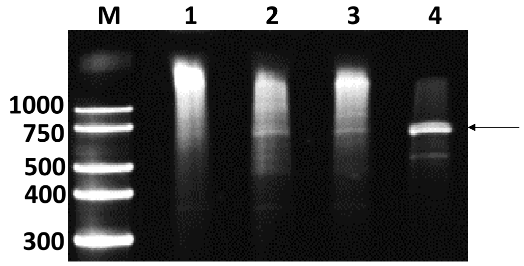


B

$$\text{Normalised ratio} = \frac{\mathit{CCMV - C19\ RNA\ copy\ number}}{\mathit{In\ vitro\ transcribed\ SDM\ RNA\ copy\ number}}$$

$$\text{Normalised copy number} = \frac{\mathit{Copy\ number\ from\ standard\ curve}}{\mathit{Normalised\ ratio}}$$

$$\text{Percentage of SDM RNAs in total RNAs} = \frac{\mathit{Normalised\ copy\ number}}{\mathit{Normalised\ CCMV{-}C19\ copy\ number}} \times 100\ \%$$

**Figure S6:** (A) Total nucleic acids extracted from VLPs were analysed on 6 % denaturing urea polyacrylamide gel. M: Thermo Fisher Scientific Century™-Plus RNA Markers. Lane 1: Qβ VLP. Lane 2: Qβ 1P-C19. Lane 3: Qβ 2P-C19. Lane 4: CCMV-C19. Arrow indicates the SDM RNA. A total of 150 ng RNA was loaded into each well. (B) Equations for normalisation of copy number.

**Table S1. Stability study: Analysis of RNAs from one-month old VLP-based SARS-CoV-2 positive controls.** Total nucleic acids include carrier RNA.

| | **Qβ 1P-C19** | **Qβ 2P-C19** | **CCMV-C19** |
|---|---|---|---|
| **Total nucleic acids (ng/µg VLP)** | 223 | 169 | 106 |
| **260/280** | 2.58 | 2.54 | 2.89 |
| **260/230** | 2.74 | 5.56 | 3.19 |

**Table S2.** Sequences of ddPCR primers and probes used in clinical settings.

| **Primer/probe** | **Sequence (5’- 3’)** |
|---|---|
| N1-F | GAC CCC AAA ATC AGC GAA AT |
| N1-R | TCT GGT TAC TGC CAG TTG AAT CTG |
| N1-P | **FAM/ZEN** - ACC CCG CAT TAC GTT TGG TGG ACC - **IBFQ** |
| N2-F | TTA CAA ACA TTG GCC GCA AA |
| N2-R | GCG CGA CAT TCC GAA GAA |
| N2-P | **FAM/ZEN** - ACA ATT TGC CCC CAG CGC TTC AG - **IBFQ** |
| RP-F | GATTTGGACCTGCGAGCG |
| RP-R | GCGGCTGTCTCCACAAGT |
| RP-P | **HEX/ZEN** - CTGACCTGAAGGCTCT- **IBFQ** |

*F refers to forward; R refers to reverse; P refers to probe

**Table S3**. Calculation of SDM RNA molecule encapsidated in VLPs.

| | **Qβ 1P-C19** | **Qβ 2P-C19** | **CCMV-C19** |
|---|---|---|---|
| **Protein concentration (µg*mol/g*mL)** | 2.60 x $10^{-5}$ | 2.94 x $10^{-4}$ | 2.66 x $10^{-5}$ |
| **Total RNA concentration (µg/mL)** | 7.42 | 51.63 | 16.71 |
| **Percentage of SDM RNA in total RNA (%)** | 3.98 | 3.80 | 100 |
| **SDM RNA concentration (µg*mol/g*mL)** | 1.48 x $10^{-6}$ | 9.86 x $10^{-6}$ | 8.40 x $10^{-5}$ |
| **Number of SDM RNA/particle** | 0.06 | 0.03 | 3.15 |
